# Analysis of signs and symptoms of SARS-CoV-2 virus infection considering different waves using Machine Learning

**DOI:** 10.1101/2024.02.12.24302722

**Authors:** Felipe C. Ulrichsen, Alexandre C. Sena, Luís Cristóvao Porto, Karla Figueiredo

## Abstract

In March 2020, the World Health Organization declared a world pandemic of COVID-19, which can manifest in humans as a consequence of virus infection of SARS-CoV-2. On this context, this work uses Data Mining and Machine Learning techniques for the infection diagnosis. A methodology was created to facilitate this task and can be applied in any outbreak or pandemic wave. Besides generating diagnosis models based only on signals and symptoms, the method can evaluate if there are differences in signals and symptoms between waves (or outbreaks) through explainable techniques of the machine learning models. Another aspect is identifying possible quality differences between exams, for example, Rapid Test (RT) and Reverse Transcription–Polymerase Chain Reaction (RT-PCR). The case study in this work is based on data from patients who sought care at Piquet Carneiro Polyclinic of the State University of Rio de Janeiro. In this work, the results obtained with the tests were used to diagnose symptomatic infection of the SARS-CoV-2 virus, based on related signals and symptoms, and the date of the initial of these signals and symptoms. Using the Random Forrest model, it was possible to achieve the result of up to 76% sensitivity, 86% specificity, and 79% accuracy in the results of tests in one contagion wave of the SARS-CoV-2 virus. Moreover, differences were found in signals and symptoms between contagion waves, in addition to the observation that exams *RT-PCR* and *RT Antigen tests* are more reliable than *RT antibody test*.

## 1 Introduction

COVID-19, Coronavirus Disease 2019, which can manifest in humans as a result of an infection caused by the SARS-CoV-2 virus, had the first cases reported in Wuhan, Hubei Province, China [1]. In March 2020, the WHO (World Health Organization) declared the COVID-19 pandemic. The ease of contagion combined with the frequent mutations of the SARS-CoV-2 virus, which even occur in the glycoprotein spike (Protein S, D614G) [24], bring on the emergence of new variants. These factors combined provide new waves of contagion around the world.

To try to reduce the number of infected people, it was necessary to test, especially people with symptoms or who had contact with people infected with the virus.

Moreover, in the absence of tests, many doctors evaluate the patient’s signs and symptoms, which are clinical manifestations perceived by the patient and are fundamental in assessing diseases. Thus, the analysis and study of signs and symptoms are essential to improve the quality and speed of diagnosis [Zhang et al. 2020] and, consequently, treatment.

On the other hand, Machine Learning (ML) is one of the areas of Artificial Intelligence (AI) that has been applied in various sectors of society, including health, since the middle of the last century [28]. Its use has greatly intensified in this area, due to the digital storage of patient data. Today its importance is recognized, with innovative perspectives in several areas of health [29], [32], [31] and [33].

Thus, Machine Learning has many advantages when applied to the health area, manipulating a high volume of variables, in a safe and reproducible way, in a much shorter period of time than a human being [34], [35], [36] and [33]. However, many of the algorithms used in the ML area are considered black boxes, making it difficult for the health area to accept the results achieved.

In this context, the objective of this work is to investigate and evaluate methodologies and models based on ML for the diagnosis of COVID-19, based only on signs and symptoms of patients, to assist health professionals during outbreaks or pandemic. The main contributions of this paper are: (i) evaluate the use of ML techniques to infer the diagnosis of COVID-19 considering different waves of contagion; (ii) increase the quality and explainability of COVID-19 diagnoses, to help healthcare professionals decide the best treatment for patients; (iii) indicate the most prevalent signs and symptoms in the different waves of contagion; (iv) evaluate and identify the variation of signs and symptoms in the different waves of contagion.

The rest of this document is organized as follows. The following section presents the related work. Materials and methods with the methodology used in this paper to evaluate signs and symptoms in different periods and tests used during the pandemic of COVID-19 are presented in Section 3. In turn, Section 4 describes the Case Study and finally, Section 5 concludes and presents prospects for future works.

## 2 Related Work

In a study conducted in Jordan [47], an online form was used to collect data for developing a diagnostic tool for COVID-19 using Multi-Layer Perceptron (MLP) and Support Vector machine (SVM). The attributes used in the study were signs and symptoms, gender and age. The study also used X-ray images in the inference which provided an accuracy above 90% for both models.

Another study, carried out in England [48], used data from more than one million participants who took part in the REACT-1 survey (REal-time Assessment of Community Transmission-1) on SARS-CoV-2 infection. The data used were: symptoms of the patients, results of the Reverse Transcription Polymerase Chain Reaction (RT-PCR) tests and results of the genetic analysis of the virus SARS-CoV-2, which were divided into two groups, anyone who was infected with the virus SARS-CoV-2 wild-type, and the other group with those infected with the B.1.1.7 (Alpha) variant of SARS-CoV-2. The LASSO algorithm was used to perform the analyses. The study obtained 72% of sensitivity and 64% of specificity for the first group (wild SARS-CoV-2 virus) and 74% of sensitivity and 64% specificity for the group with the Alpha variant. Initially, tests were performed with 26 different symptoms, which were later reduced to the seven following symptoms that provided the best results: anosmia, ageusia, fever, cough, chills, lack of appetite and myalgia.

In turn, the study carried out in the United Kingdom [49], used a cell phone application for users to inform the signs and symptoms after the third day of the first symptom and the presence of pre-existing diseases. The Hierarchical Gaussian model, Bayesian framework and Logistic Regression were used. There was a division into groups: health professionals or not, gender, age, body mass index and date of onset of symptoms. There was no data equalization, but an attempt was made to reduce the imbalance between negatives and positives in the database using bootstrapping, where the percentage of positives increased from 2% to 5%. The best result was obtained in the groups of health workers using the hierarchical Gaussian model, achieving a sensitivity of 76%. Although, in this study, the most relevant symptoms varied between groups, in the younger people group, for example, only anosmia and chest pain were significant for the positive diagnosis for COVID-19.

A similar study in England [50] used data from the United Kingdom and the United States of America that were obtained through a cell phone application in which patients reported symptoms, BMI (body mass index), sex, pre-existing diseases, demographic data and the result of the RT-PCR test, in the period from March 24, 2020 to April 21 2020. The Logistic Regression algorithm was used to make the inferences and the data were not equalized. Instead, the inputs were divided into groups by sex, age and BMI. The study had a mean sensitivity of 65% and mean specificity of 78% for UK data and mean sensitivity of 66% and mean specificity of 83% for USA data. The following symptoms were associated with a positive result: Anosmia, lack of appetite, fatigue, fever, cough, diarrhea, delirium, hoarseness, breathing difficulty, abdominal pain and neck pain.

On the other hand, the work presented in [52] developed a Machine Learning model based on signs and symptoms, age, gender and whether there was contact with confirmed cases of COVID-19. The Israeli Ministry of Public Health provided data with RT-PCR results collected from March 2, 2020 to April 7, 2020. There was no equalization of data between positive and negative. The gradient-boosting model was used, with results that reached 0.90 in the area under the ROC (AUC) curve of sensitivity x specificity. Using Shapley, a game-theoretic explainable technique, it was concluded that the most significant attributes for the diagnosis were: cough, fever, whether a person had contact with a confirmed case of COVID-19, if you are a man, if you are over 60 years old, headache, sore throat and dyspnea.

Still, in the context of COVID-19 diagnosis, a study carried out in the State of Rio de Janeiro [33] focused on identifying the underreporting of COVID-19 cases. Data were obtained through an electronic form with self-reported signs and symptoms and onset of symptoms of COVID-19. To infer whether or not a given respondent had COVID-19, models were developed based on Machine Learning. The best model indicated accuracy just above 60%. This model was used to identify respondents who were possibly sick and not tested, being considered underreported cases.

Differently from the works presented in this section, the work proposed in this article analyzes the signs and symptoms by waves and types of tests used to diagnose COVID-19. To highlight the differences, a comparison of the work proposed in this article and the studies presented in this section can be seen in Table 1.

**Table 1.**
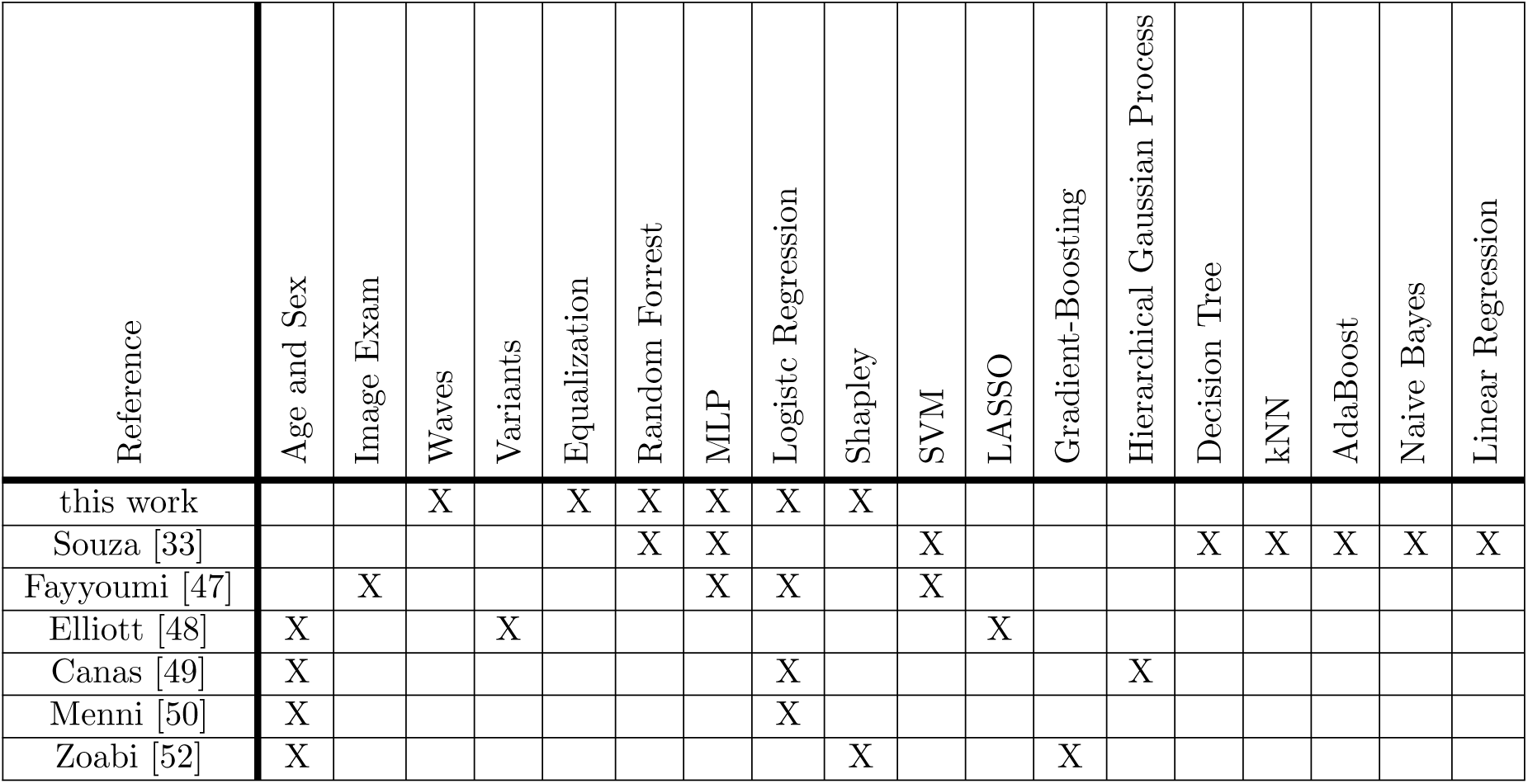
Comparison of proposed work with related work.

As can be clearly seen, differently from the other works, only signs and symptoms described by health professionals and the COVID-19 test results are used as attributes Moreover, data were divided and analyzed in waves to assess the impact on the model.

This work is also the only one that equalizes the data between positive and negative for COVID-19 tests. Although the real world is not equalized, evaluating models with balanced data is important to eliminate possibly biased results. In addition, this work also analyzes the influence of signs and symptoms on the result generated by the machine learning model through explainable and Shapley techniques.

## 3 Materials and methods

The context of data manipulation by Machine Learning algorithms normally inserts it in the Data Mining process for extracting information from databases. Thus, based on the process applied in Data Mining, the data must be initially evaluated, verifying the need to apply data pre-processing techniques. In general, it is necessary to organize the data identify outliers and missing data, normalize and equalize databases and, finally, select variables, so that the algorithms can make the most of the data and present better results [41] [42] [43].

An overview of the methodology adopted in this work can be seen in Figure 1. The process can be divided into three phases: data acquisition, pre-processing and application and, finally, analysis of classification results obtained by Machine Learning Initially, the data corresponding to the patient’s signs and symptoms and the results of the COVID-19 tests are obtained through health information systems (Figure 1.(1)) or by any electronic form (Figure 1.(2)). In turn, the preprocessing step starts with the combination of the corresponding data identified in the databases (signs and symptoms + test results), if they are stored in different files (Figure 1.(3)). It is essential to highlight that the data does not need to be identified, that is, personal or sensitive data do not need to be available. Then, when applicable, quantitative data is normalized, Figure 1.(4). Qualitative attributes, sortable or not, must be transformed into numeric data. In the case of this work, the attributes are categorical and dichotomous, being therefore transformed into zero (0) or one (1). Still, during the data preprocessing stage, the data are separated by contagion waves, Figure 1.(5), that is, the data are divided based on the start and end dates of the outbreaks, and have their bases balanced with respect to the output attribute (the number of patients with COVID-19 test results positive and negative). The next step is the selection of attributes, Figure 1.(6), with the application of filter-type techniques, selecting the most relevant variables for the Machine Learning algorithms, aiming to maximize the classification accuracy.

**Fig 1.**
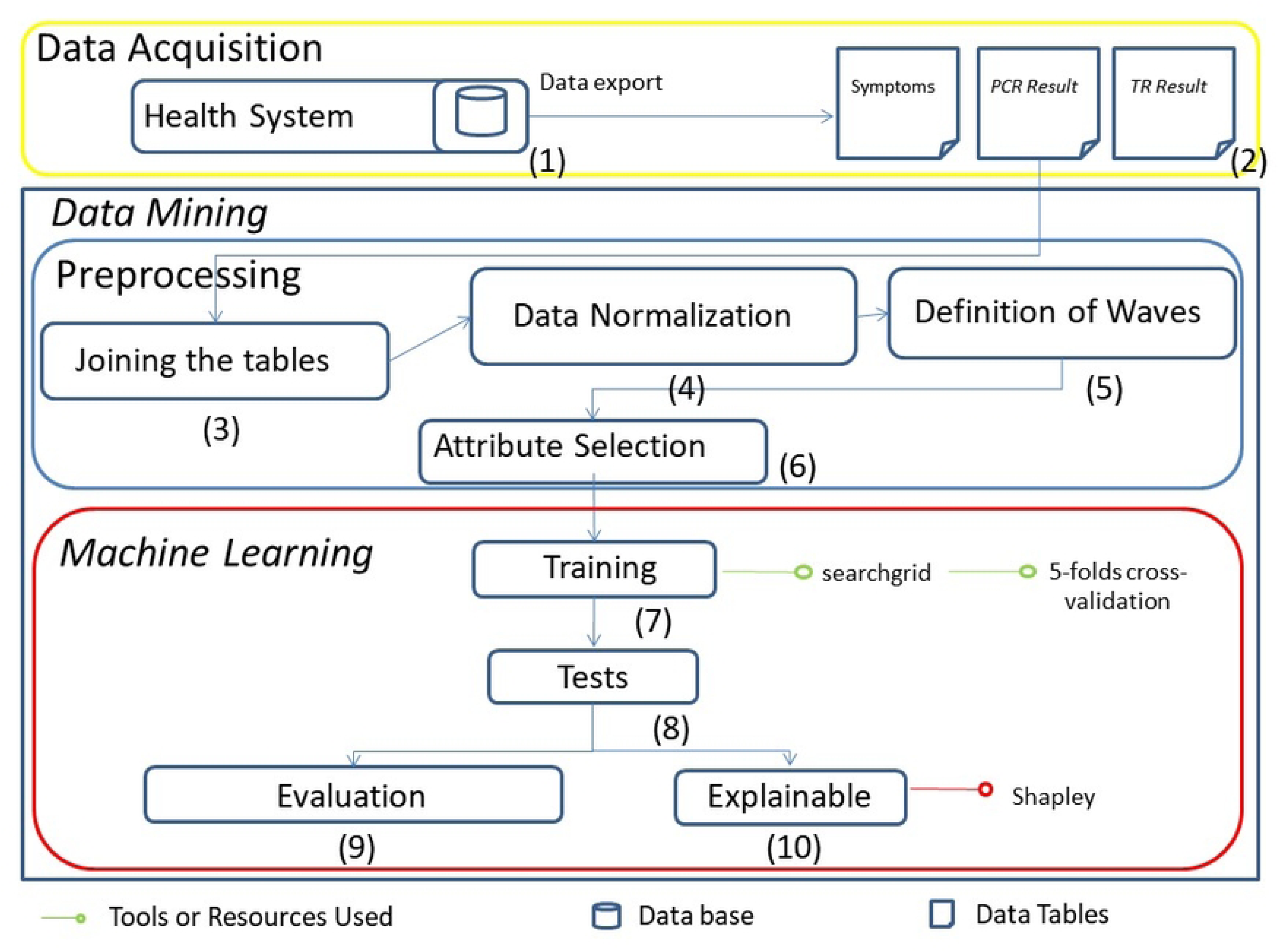

After data preprocessing, stage 3 starts with the training of the Machine Learning algorithms, Figure 1.(7). Each model is proposed based on the definition of its hyperparameters, followed by the evaluation of its results through cross-validation, which can be based on one or more metrics (in general, the averages of accuracy, recall, precision and f1 and the ROC/AUC) obtained with the *n* models during cross-validation The best average cross-validation results identify the most suitable algorithms/models. Once the models are selected, the testing phase is started (Figure 1.(8)), which is evaluated (Figure 1.(9)) by the same metrics used in the validation and, in general, plus the confusion matrix. Finally, in order to understand the importance of each attribute in the obtained result, the phase of understanding the results (Explainable), Figure 1.(10), is carried out, using, for example, the method from Shapley.

Thus, five algorithms were selected, four for the COVID-19 prediction, focusing on the specificities of each wave, and one for the Explainable analysis: Random Forrest [9], Multi-Layer Perceptron [10], XGBoost [45], Logistic Regression [46] and the Shapley Additive Explanation for the input importance analysis. This set of algorithms was chosen to investigate the database’ linear and nonlinear characteristics and explore each algorithm separately and combined (ensemble).

The Random Forest (RF) algorithm is an ensemble decision tree algorithm that generates many classifiers and combines their results. The algorithm uses bootstrap aggregating or bagging, by randomly selecting records, with replacement, to be used in the construction of each tree, reducing variance without harming bias [9]. Breiman [8] observed that there was a deep correlation between decision trees, and to reduce this effect, a random selection of attributes that may be available for the construction of each tree, or even each branch of the tree, was implemented, decreasing the correlation between the decision trees.

The XGBoost was developed primarily to increase the performance and speed at which small decision trees are created to reduce the errors of previous ones, using gradient-boosted. In the case of the gradient-based “boostin” algorithm, the errors made by previous trees as they are created are minimized by the decreasing gradient algorithm, and this algorithm has been proposed by Tianqi Chen [11] and applied by many other developers. XGBoost or Extreme Gradient Boosting combines software and hardware optimization techniques to produce superior results. This has the benefit of improving the algorithm by tuning the model for better performance. The XGBoost can choose among three gradient boosting techniques: gradient boosting, regularized gradient boosting and Stochastic Boosting. It is effective in reducing computation time provided by optimal use of memory resources. In addition, the algorithm can handle missing values, supports parallel structures when building trees, and has the unique quality of boosting performance by adding data to already trained models (Continuous Training) [12] [13].

The Multi-Layer Perceptron (MLP) is an algorithm inspired by biological neurons. It is based on constructing Neural Networks (NN)(interconnecting neurons between layers) that learn to map input data relationships to output data, in supervised problems, by adjusting synaptic weights based on errors identified during the learning process [53].

Logistic regression is a simple statistical technique widely used in many areas. It aims to generate, from input data (whether numerical or nominal), a linear model that allows the prediction of values defined by a categorical variable, usually binary [14].

Lundberg and Lee [19] included Shapley Additive Explanations as an approach to explain the output of machine learning algorithms, enabling model interpretation. The Shapley algorithm’s essence is to measure each variable’s contribution to the final result This algorithm category has become important for models that are not intrinsically explainable.

## 4 Results and Discussion

This section presents and evaluates the results considering the diagnostic investigation from the perspective of different waves of the COVID-19 pandemic, and the qualification of the tests used.

### Database

All data used in this research refer to the COVID-19 tests carried out at the Piquet Carneiro Polyclinic, which is part of the health complex of the State University of Rio de Janeiro.

This study was conducted in accordance with ethical principles outlined in the Declaration of Helsinki and was approved by the Pedro Ernesto University Hospital Ethical Committee (CAAE: 30135320.0.0000.5259).

The symptoms freely described on the form by the patients were unified (e.g., “cephalea and headache”), based on the analysis of three evaluators. To perform the diagnoses, the following laboratory tests were considered: RT-PCR, Rapid Antibody Test (RT-antibody), and Rapid Antigen Test (RT-antigen).

Serologic antibody tests are used to identify infection (IgM) or immunity (IgG) to COVID-19. Results can be positive for immunoglobulin G and M antibodies from days 4-5 of symptom onset. In general, 70% of patients show IgM type antibodies within 8-14 days of symptom onset and 98% of IgG after several weeks, but the duration of this immune response is not rigid and may vary from person to person [38]. In turn, nasal RT-PCR should be performed within 3-7 days of the onset of symptoms [39]. On the other hand, the Rapid Antibody Test (RT-antibody) (a rapid chromatographic immunoassay for the qualitative detection of specific antigens of SARS-CoV-2 present in the human nasopharynx), is based on the identification of antibodies and thus should be performed after day seven from the onset of symptoms. The RT-PCR is considered the standard test for COVID-19. More recently, a new type of Rapid Test has been increasingly used, the antigen RT. The latter is considered more accurate than the RT-antibody. As can be seen, some tests may be more or less appropriate depending on the time the test is performed and the onset of signs and symptoms.

From Figures 2 and 3, the start and end dates of the first, second and third waves were defined. These figures indicate the moving average of patients notified by the Rio de Janeiro state health system. Thus, the start and end dates of the first wave are 03/18/2020 and 06/18/2020 (Figure 2). Similarly, based on the analysis of those notified by the Rio de Janeiro State health system (Figures 2 and3), we obtained the start and end dates for the second wave, 10/18/2020 and 2/18/2021, respectively, and the dates 12/25/2021 and 2/25/2022, for the start and end for the third wave (Figure 3). Possible waves are indicated with red rectangles in Figure 3 that presents the chart of confirmed cases of symptomatic infection by the SARS-CoV-2 virus by date of onset of symptoms in the city of Rio de Janeiro. The light blue line represents the 7-day moving average^1^

**Fig 2.**
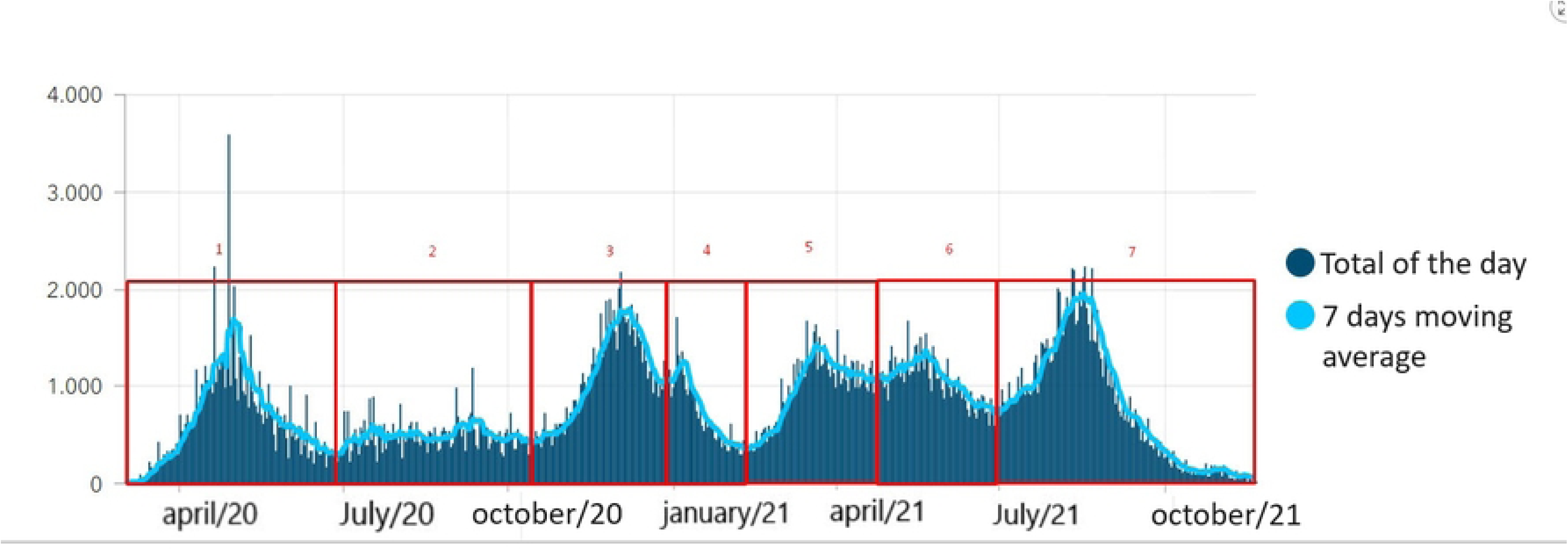

**Fig 3.**
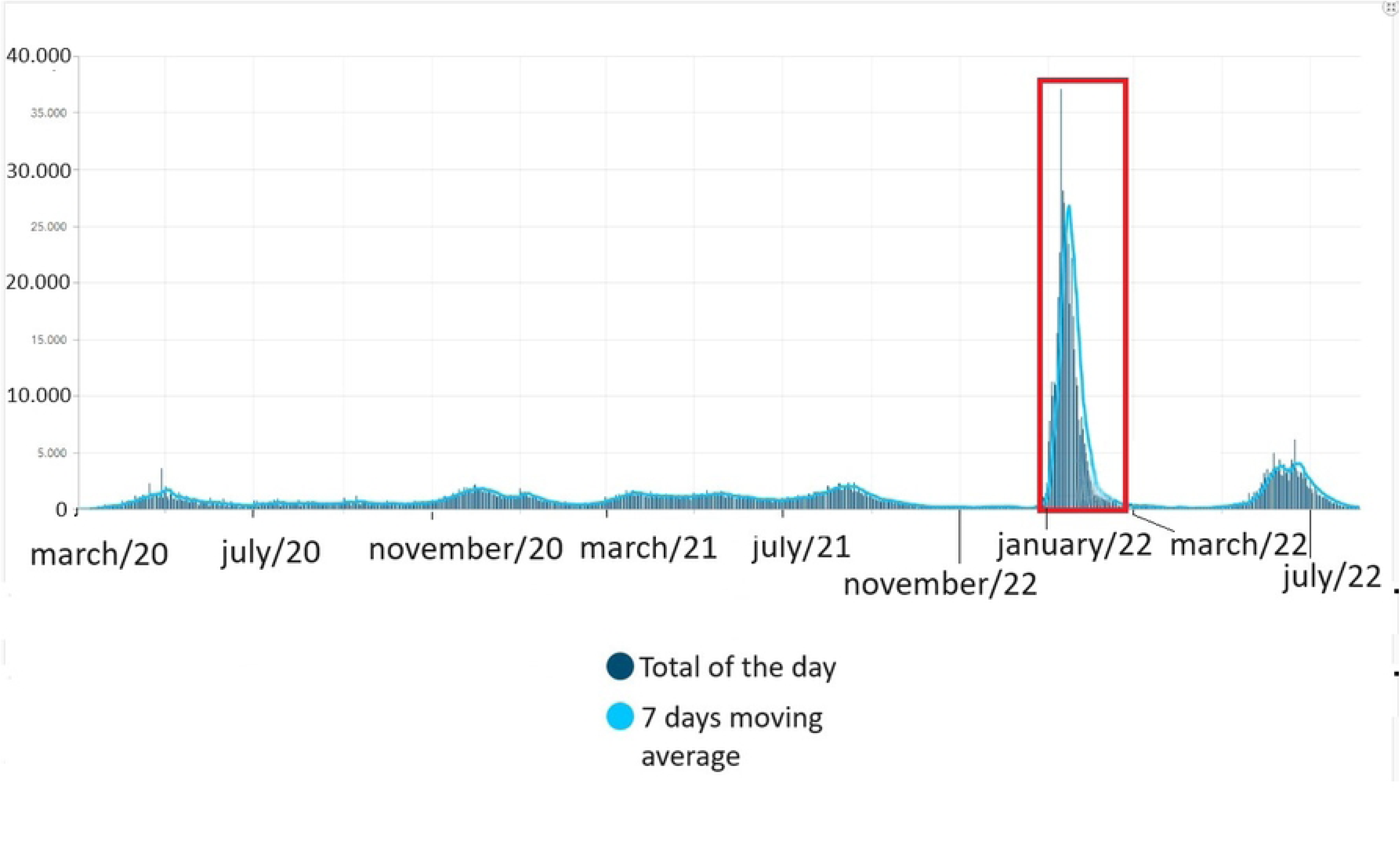

After evaluation by health specialists, 19 signs and symptoms, with the highest prevalence, were selected for 1st and 2nd waves and 19 signs and symptoms for 3rd wave (not exactly the same signs and symptoms). At this first stage, a decision was made to consider these 19 signs and symptoms more broadly, including those with low representativeness in the database. This approach allows the variable selection step and the machine learning algorithms to purge signs and symptoms that do not contribute to discriminating the diagnosis in each of the three waves.

Figure 4 shows the prevalence of the 19 signs and symptoms in patients with negative and positive results, according to the RT-PCR test, considered from the first and second waves. On the other hand, Figure 5 presents a bar chart depicting the primary signs and symptoms reported by patients during the third wave, all of whom underwent the Rapid Antigen Test at PPC, with samples collected by nasal swab.

**Fig 4.**
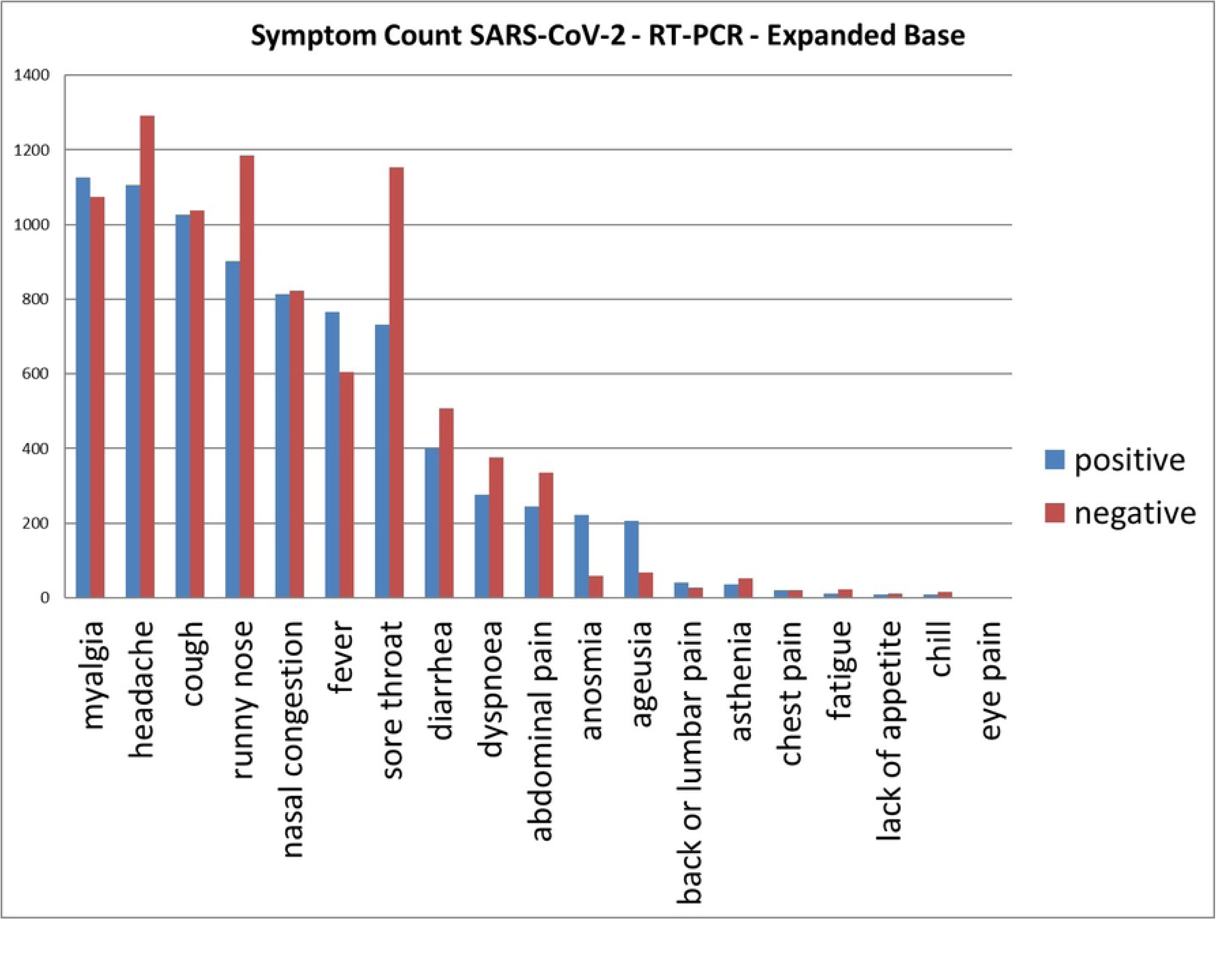

**Fig 5.**
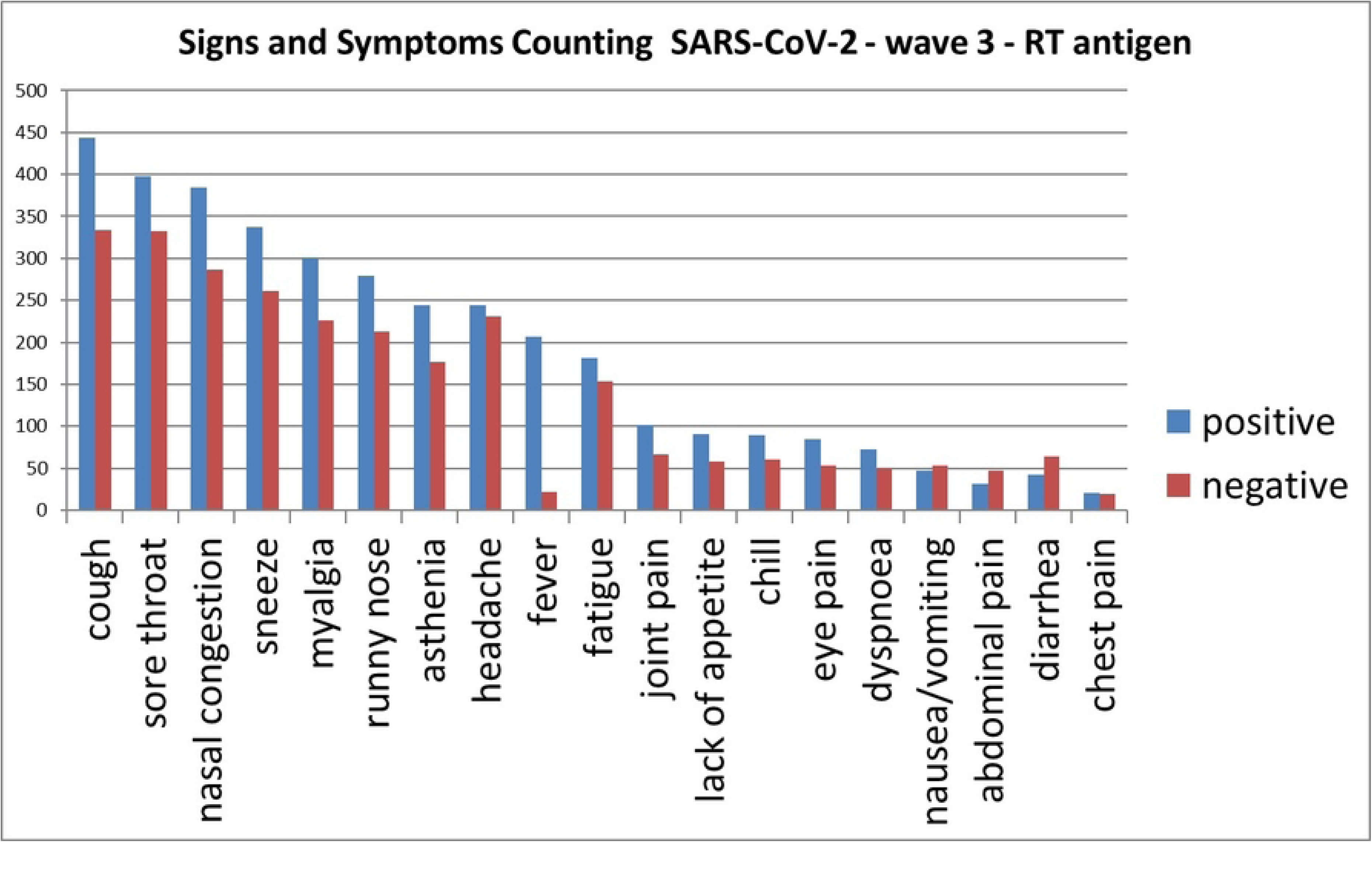

### Preprocessing Database

It is important to highlight that, for a detailed analysis, instead of only analyzing data from the first, second, and third waves, data were divided according to the different waves and types of tests (RT-PCR, RT-Antigen, and RT-antibody) performed to diagnose the patient as positive or negative.

This division resulted in 10 datasets that can be seen in the list below and the first column of Table 2, and also in the first column of most of the following tables (i.e. column with the label Data group).

1. RT-PCR (1*^st^* wave + 2*^nd^* wave)
2. RT-PCR 1*^st^* wave
3. RT-PCR 2*^nd^* wave
4. RT-antigens 3*^rd^* wave
5. RT-antibody (1*^st^* wave + 2*^nd^*wave)
6. RT-antibody 1*^st^* wave
7. RT-antibody 2*^nd^* wave
8. RT-PCR + RT-antibody (1*^st^* wave + 2*^nd^* wave)
9. RT-PCR + RT-antibody 1*^st^* wave
10. RT-PCR + RT-antibody 2*^nd^* wave

**Table 2.** Number of records for each database: using approximately 90% for training and validation, and 10% for testing.

| Data group | Training and validation | Test |
| --- | --- | --- |
| 1 - RT-PCR (1<sup>st</sup> wave + 2<sup>nd</sup> wave) | 2436 | 272 |
| 2 - RT-PCR 1<sup>st</sup> wave | 1228 | 138 |
| 3 - RT-PCR 2<sup>nd</sup> wave | 839 | 93 |
| 4 - RT-antigens 3<sup>rd</sup> wave | 1080 | 120 |
| 5 - RT-antibody (1<sup>st</sup> wave + 2<sup>nd</sup> wave) | 1990 | 222 |
| 6 - RT-antibody 1<sup>st</sup> wave | 1748 | 196 |
| 7 - RT-antibody 2<sup>nd</sup> wave | 67 | 9 |
| 8 - RT-PCR + RT-antibody (1<sup>st</sup> wave + 2<sup>nd</sup> wave) | 4426 | 494 |
| 9 - RT-PCR + RT-antibody 1<sup>st</sup> wave | 2926 | 332 |
| 10 - RT-PCR + RT-antibody 2<sup>nd</sup> wave | 769 | 87 |

The quantitative records of patients with signs and symptoms related to the type of test and its results (negative and positive) for the first, second and third waves were unbalanced. Therefore, for the models to have a better performance, without bias influence, data equalization was performed, i.e., based on the number of positives, entries with negative results were randomly chosen, making a balance by undersampling the class with the major number of records.

The data from the first, second and third waves, that indicated symptoms, were defined as input attributes in dichotomous format (zero and one). Thus, the symptoms were defined as “1” for the patient who indicated that he had the symptom and “0” otherwise. In addition, the diagnosis column was also categorized as “1” if the test result (RT-PCR, RT-antibody or RT-antigen) was positive and “0” when negative for SARS-CoV-2 infection.

After that, approximately 10% of positive and negative cases were separated from the balanced base to compose the test base, and the rest was left for model training and validation. This procedure was performed on all bases, and Table 2 shows the amount of data for each phase (i.e. Training and Validation or Test).

### Model Evaluation and Attribute Selection

A detailed evaluation of attributes was carried out separately for each base. A sensitivity analysis type evaluation was performed, which led the models to the best results; such analysis consists of removing each of the signs and symptoms and evaluating the improvement in validation results, (i.e., after the improvement evaluation by removing one attribute, we proceeded with other evaluations verifying the removal of a second attribute). This evaluation only included data groups 2, 3, and 4.

This evaluation considered each algorithm indicated (Multi-Layer Perceptron, Random Forest and Linear Regression) using the hyperparameters presented in Table 3 which were applied to all groups (from 1 to 10). Through exploratory analysis, variable selection, and the search for optimal hyperparameters (using a search grid) (Table 3), we identified the signs and symptoms that most effectively enhance the models, leading to better results.

**Table 3.** Parameter settings used in SearchGrid for MLP, RF and LR.

|  |  |  |
| --- | --- | --- |
| **MLP** | hidden_layer_sizes | 5, 10, 15, 20, 30, 40, 50, 60, 70, 80, 90, 100 |
|  | activation | relu, tanh |
|  | solver | adam, sgd, lbfgs |
|  | alpha | 0.001, 0.05 |
|  | learning_rate | constant, adaptive |
|  | nesterovs_momentum | True, False |
|  | beta_1 | 0.95 |
|  | beta_2 | 0.999 |
|  | learning_rate_init | 0.0001, 0.005, 0.001, 0.05 |
|  | momentum | 0.9, 0.95 |
| **RF** | hidden_layer_sizes | 5, 10, 15, 20, 30, 40, 50, 60, 70, 80, 90, 100 |
|  | activation | relu, tanh |
|  | solver | adam, sgd, lbfgs |
|  | alpha | 0.001, 0.05 |
|  | learning_rate | constant, adaptive |
|  | nesterovs_momentum | True, False |
|  | beta_1 | 0.95 |
|  | beta_2 | 0.999 |
|  | learning_rate_init | 0.0001, 0.005, 0.001, 0.05 |
|  | momentum | 0.9, 0.95 |
| **LR** | tol | 0.5, 0.001 |
|  | C | 1.0, 2.0, 3.0 |
|  | fit_intercept | True, False |
|  | class_weight | None |
|  | solver | lbfgs, saga |

Furthermore, this evaluation for the best attribute choice was jointly considered with searching for better hyperparameters from the cross-validation method, which divided the dataset into 5 equal parts stratified (each part has the same amount of positive and negative diagnostics). Thus, the models were built with four parts of the database for training and one part for validation. This procedure was performed five times and, at least once, one of the five parts was used as the validation set. Finally, the model, per database, with the best average accuracy was selected. The main metric for selecting the best model was accuracy, but other measures of model performance were used, such as precision, Recall, F1, AUC, sensitivity, and specificity.

Table 4 shows the attributes that produced the best results for each wave, the study was carried out for data groups 2, 3 and 4. So, after evaluating the results with the removal (individually) of each remaining attribute, it was concluded that after removing “nasal congestion” and “chest pain” in the case of the first and second wave and only “chest pain” for the third wave, removing more attributes from the databases was not appropriate.

**Table 4.** Attributes selected by sensitivity analysis with best wave validation result.

| *1<sup>st</sup> wave* | *2<sup>nd</sup> wave* | *3<sup>rd</sup> wave* |
| --- | --- | --- |
| ageusia | ageusia | asthenia |
| anosmia | anosmia | chills |
| asthenia | asthenia | headache |
| chills | chills | nasal congestion |
| headache | headache | runny nose |
| runny nose | runny nose | diarrhea |
| diarrhea | diarrhea | dyspnoea |
| dyspnoea | dyspnoea | abdominal pain |
| abdominal pain | abdominal pain | joint pain |
| back and lumbar pain | back and lumbar pain | sore throat |
| sore throat | sore throat | eye pain |
| eye pain | eye pain | sneeze |
| fatigue | fatigue | fatigue |
| fever | fever | fever |
| lack of appetite | lack of appetite | lack of appetite |
| myalgia | myalgia | myalgia |
| cough | cough | nausea or vomiting |
|  |  | cough |

In Table 4 can be seen that the signs and symptoms of the 1*^st^* and 2*^nd^* wave, with the best validation results, were the same. On the other hand, in the 3*^rd^* wave, some signs and symptoms that better helped the model classify positive or negative for SARS-CoV-2 infection were different.

Table 5 presents the best results obtained for the databases considered, with the best hyperparameters shown in Tables 6, 7 and 8. Again, each database had its hyperparameters determined by the hyperparameter search algorithm, and the criteria were based on the validation metrics.

**Table 5.** Metrics - average validation errors.

| Method | Data group | #records | Accuracy | Precision | Recall | f1 | AUC |
| --- | --- | --- | --- | --- | --- | --- | --- |
| RF | 1 | 2852 | 0.65 | 0.67 | 0.60 | 0.63 | 0.70 |
|  | 2 | 1228 | 0.62 | 0.61 | 0.53 | 0.57 | 0.66 |
|  | 3 | 839 | 0.70 | 0.70 | 0.64 | 0.67 | 0.71 |
|  | 4 | 1080 | 0.67 | 0.67 | 0.65 | 0.66 | 0.68 |
|  | 5 | 1990 | 0.64 | 0.64 | 0.53 | 0.58 | 0.65 |
|  | 6 | 1748 | 0.64 | 0.64 | 0.48 | 0.55 | 0.65 |
|  | 7 | 67 | 0.64 | 0.64 | 0.67 | 0.66 | 0.66 |
|  | 8 | 4426 | 0.65 | 0.67 | 0.59 | 0.62 | 0.68 |
|  | 9 | 2956 | 0.62 | 0.64 | 0.54 | 0.58 | 0.66 |
|  | 10 | 769 | 0.69 | 0.70 | 0.67 | 0.68 | 0.72 |
| MLP | 1 | 2852 | 0.65 | 0.67 | 0.60 | 0.63 | 0.70 |
|  | 2 | 1228 | 0.62 | 0.62 | 0.61 | 0.62 | 0.65 |
|  | 3 | 839 | 0.69 | 0.70 | 0.64 | 0.67 | 0.71 |
|  | 4 | 1080 | 0.66 | 0.66 | 0.68 | 0.66 | 0.68 |
|  | 5 | 1990 | 0.64 | 0.65 | 0.59 | 0.62 | 0.66 |
|  | 6 | 1748 | 0.61 | 0.66 | 0.47 | 0.55 | 0.65 |
|  | 7 | 67 | 0.62 | 0.66 | 0.47 | 0.55 | 0.64 |
|  | 8 | 4426 | 0.67 | 0.65 | 0.67 | 0.67 | 0.68 |
|  | 9 | 2956 | 0.68 | 0.65 | 0.68 | 0.68 | 0.65 |
|  | 10 | 769 | 0.68 | 0.66 | 0.68 | 0.68 | 0.61 |
| RL | 1 | 2852 | 0.65 | 0.65 | 0.62 | 0.62 | 0.70 |
|  | 2 | 1228 | 0.61 | 0.62 | 0.54 | 0.57 | 0.64 |
|  | 3 | 839 | 0.68 | 0.66 | 0.76 | 0.71 | 0.74 |
|  | 4 | 1080 | 0.65 | 0.66 | 0.61 | 0.64 | 0.68 |
|  | 5 | 1990 | 0.62 | 0.63 | 0.59 | 0.61 | 0.65 |
|  | 6 | 1748 | 0.61 | 0.64 | 0.51 | 0.56 | 0.64 |
|  | 7 | 67 | 0.57 | 0.65 | 0.50 | 0.53 | 0.51 |
|  | 8 | 4426 | 0.63 | 0.64 | 0.60 | 0.62 | 0.67 |
|  | 9 | 2956 | 0.61 | 0.66 | 0.43 | 0.52 | 0.64 |
|  | 10 | 769 | 0.66 | 0.67 | 0.66 | 0.66 | 0.72 |

**Table 6.** Best hyperparameters per data group - Random Forest.

| Data group | criterion | max_depth | max_features | min_samples_leaf | min_samples_split |
| --- | --- | --- | --- | --- | --- |
| 1 | entropy | 10 | 2 | 4 | 8 |
| 2 | entropy | 4 | 2 | 7 | 4 |
| 3 | gini | 10 | 4 | 7 | 2 |
| 4 | gini | 4 | 8 | 2 | 16 |
| 5 | gini | 8 | 2 | 3 | 16 |
| 6 | entropy | 8 | 3 | 6 | 2 |
| 7 | entropy | 10 | 2 | 8 | 16 |
| 8 | gini | 7 | 3 | 7 | 100 |
| 9 | entropy | 10 | 2 | 8 | 100 |
| 10 | gini | 6 | 3 | 8 | 100 |

**Table 7.**
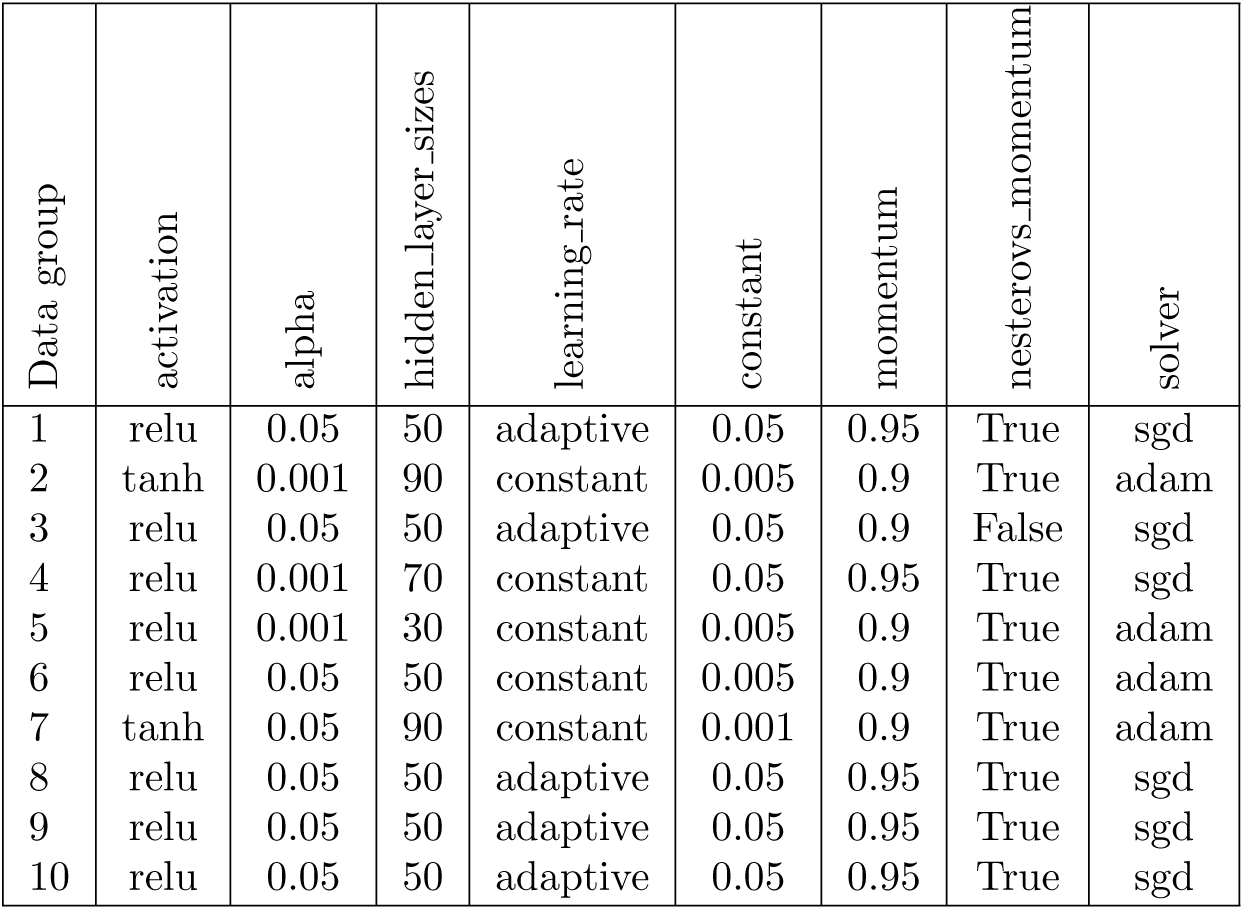
Best hyperparameters per data group - Multi-Layer Perceptron.

**Table 8.** Best hyperparameters per data group - Logistic Regression.

| Data group | C | fit_intercept | solver | tol |
| --- | --- | --- | --- | --- |
| 1 | 1.0 | True | lbfgs | 0.001 |
| 2 | 1.0 | True | saga | 0.5 |
| 3 | 1.0 | True | lbfgs | 0.5 |
| 4 | 1.0 | True | lbfgs | 0.001 |
| 5 | 2.0 | True | lbfgs | 0.5 |
| 6 | 3.0 | True | saga | 0.5 |
| 7 | 1.0 | False | saga | 0.5 |
| 8 | 1.0 | True | lbfgs | 0.001 |
| 9 | 1.0 | True | saga | 0.5 |
| 10 | 1.0 | True | lbfgs | 0.001 |

Observing Table 5 and evaluating the results for all metrics loosely (without considering the significance of the proximity between the results), it can be mentioned that:

- for data group 1, RT-PCR (1*^st^* wave + 2*^nd^*wave), the RF and MLP algorithms obtained equal measures for all metrics evaluated, and RL obtained almost equal values except for precision, recall and f1.

- the MLP algorithm showed slightly better results for the RT-PCR 1*^st^* wave (data group 2) than RF and RL.

- the RF and MLP algorithms also remained almost identical for the RT-PCR 2*^nd^*wave database (data group 3). However, the results for the metrics recall, f1 and AUC were superior when using the RL algorithm.

- for RT-antigens for the third wave (data group 4), the RF and MLP algorithms had nearly identical metrics and RL showed slightly lower results.

- overall, the RL algorithm outperformed the RF and RL algorithms for all

RT-antibody bases (data groups 5,6 and 7), with a highlight to the 64% accuracy for the base relative to the patients of the second wave (data group 7). This result is out of line, but the very small amount of data from this base should be noted.

- the MLP algorithm (considering all metrics) performed better for the case of the bases with the total number of patients tested by RT-PCR and RT-antibody were aggregated (data group 8 and 9), except for the case of the second wave (data group 10) where RF had the best result.

Applying the Mann-Whitney test on the values of the average accuracies, the following p-values *>* 0.05 were obtained for the comparison of the results obtained between MLP and RF, MLP and RL and RF and RL: p-value= 1, p-value= 0.1211 and p-value= 0.1415, respectively. These values mean that the results are not significantly different.

Applying the Mann-Whitney test on the mean values of the precision metrics, the following p-values *>* 0.05 were obtained for the comparison of the results obtained between MLP and RF, MLP and RL and RF and RL: p-value= 0.9362, p-value= 0.2713 and p-value= 0.4472, respectively. These values mean that the results, from the point of view of the precision metric, are not significantly different either.

Applying the Mann-Whitney test on the mean values of the algorithms recall metrics, the following p-values *>* 0.05 were obtained for the comparison of the results obtained between MLP and RF, MLP and RL and RF and RL: p-value= 0.4065, p-value= 0.3628 and p-value= 0.7641, respectively. Again, these results mean that from the point of view of the recall metric, they are not significantly different either.

In order to highlight the results, Table 9 presents the best average results of validation accuracy (considering the exhaustive combinations of hyperparameters indicated in Table 3), for all algorithms evaluated applied to all 10 data groups. Although the metric chosen for selecting the best model was accuracy, the metrics recall f1 and AUC were also calculated and would serve, in this order, to break the tie in case of very close accuracy values.

**Table 9.** Validation with medium accuracy (k-fold=5) with the best evaluated hyperparameter set.

| Data group | MLP | RF | RL |
| --- | --- | --- | --- |
| 1 | 0.65 | 0.65 | 0.65 |
| 2 | 0.62 | 0.62 | 0.61 |
| 3 | 0.69 | 0.70 | 0.68 |
| 4 | 0.66 | 0.67 | 0.63 |
| 5 | 0.64 | 0.64 | 0.62 |
| 6 | 0.61 | 0.64 | 0.61 |
| 7 | 0.62 | 0.64 | 0.57 |
| 8 | 0.67 | 0.65 | 0.63 |
| 9 | 0.68 | 0.62 | 0.61 |
| 10 | 0.68 | 0.69 | 0.66 |

Applying the Mann-Whitney test, it obtains p-values *<* 0.05 for the comparison of the results obtained by MLP and RF (p-value= 0.009) and by MLP and RL (p-value= 0.002), but the test obtains a result p-value*>* 0.05 (p-value= 0.110) for RF and RL. This means that the results of the MLP are significantly lower and can be considered better than those obtained by RF and RL; the same cannot be said for the results of RF and RL, i.e., they are equivalent results.

Considering the accuracy, the evaluation of the base RT-PCR 2*^nd^* wave (data group 3), in general, obtained the best diagnostic performance, also reflected in the RT-PCR (1*^st^* wave + 2*^nd^* wave) database (data group 1). These results may indicate that these signs and symptoms were more decisive in discriminating patients positive or negative for SARS-COV-2 infection, associated with the test being more reliable in the results generated, i.e., the positive or negative labels are more reliable than the results produced by other tests. On the other hand, the failures of the RT-antibody, reducing the confidence in the results, helped reduce the performance of the models for the bases associated with this test.

Nevertheless, since it is necessary to choose one of the algorithms, we opted for the RF algorithm, which outperforms MLP and RL on most bases for the average accuracy results (Table 9).

### Evaluation of Test Results

In this section, the evaluation of the results of the various tests performed, using the ML models in the different bases, will be presented.

Observing Table 10, it can be seen that the Random Forrest model achieved better metrics: 79% in accuracy using the RT-PCR 2*^nd^* wave database (data group 3). Also noteworthy is the 85% in the precision metric, as can be seen in Table 10. It is also important to highlight that the model achieved 76% of Sensitivity and 82% of Specificity, indicating that in 76% of the positive cases for SARS-COV-2 infection, the model gets it right. The results using the RT-antibody databases (data groups 5,6 and 7) confirmed the worst performance for all models, following what the results of validation (Table 9) had already indicated.

**Table 10.**
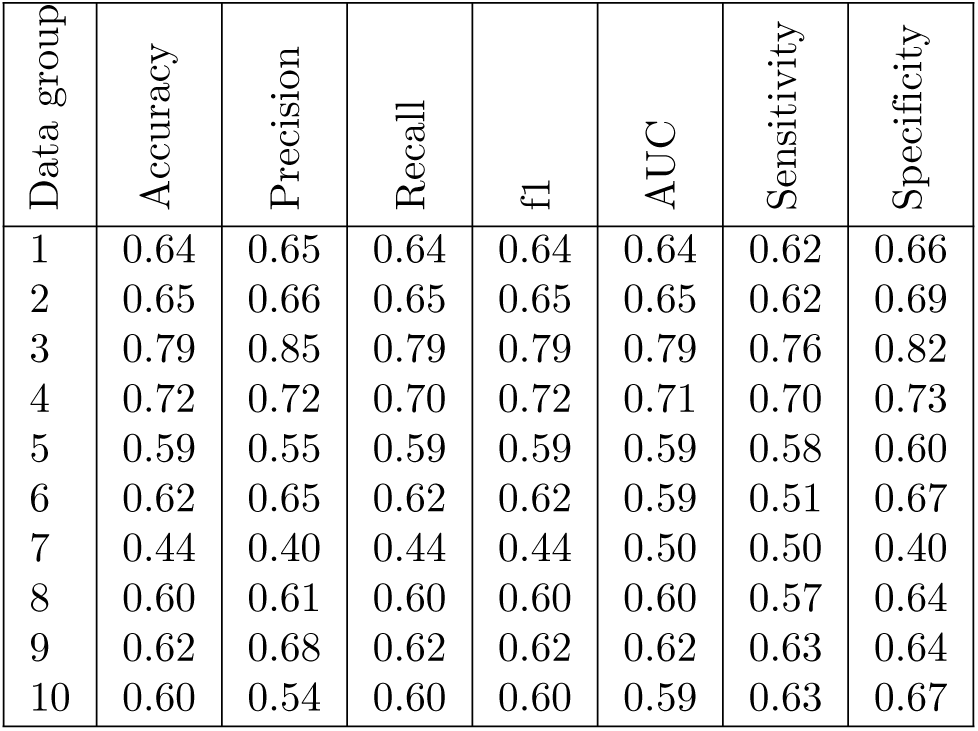
Metrics for the results with the test set - Random Forrest.

| Data group | Accuracy | Precision | Recall | f1 | AUC | Sensitivity | Specificity |
| --- | --- | --- | --- | --- | --- | --- | --- |
| 1 | 0.64 | 0.65 | 0.64 | 0.64 | 0.64 | 0.62 | 0.66 |
| 2 | 0.65 | 0.66 | 0.65 | 0.65 | 0.65 | 0.62 | 0.69 |
| 3 | 0.79 | 0.85 | 0.79 | 0.79 | 0.79 | 0.76 | 0.82 |
| 4 | 0.72 | 0.72 | 0.70 | 0.72 | 0.71 | 0.70 | 0.73 |
| 5 | 0.59 | 0.55 | 0.59 | 0.59 | 0.59 | 0.58 | 0.60 |
| 6 | 0.62 | 0.65 | 0.62 | 0.62 | 0.59 | 0.51 | 0.67 |
| 7 | 0.44 | 0.40 | 0.44 | 0.44 | 0.50 | 0.50 | 0.40 |
| 8 | 0.60 | 0.61 | 0.60 | 0.60 | 0.60 | 0.57 | 0.64 |
| 9 | 0.62 | 0.68 | 0.62 | 0.62 | 0.62 | 0.63 | 0.64 |
| 10 | 0.60 | 0.54 | 0.60 | 0.60 | 0.59 | 0.63 | 0.67 |

Among the results of RT-antibody, it is important to highlight the worst performance for the RT-antibody 2*^nd^* wave (data group 7), which certainly had its result also hampered by the limited amount of data. The model could not predict the results in a minimally satisfactory way.

### Test with data outside the Confidence Interval

Therefore, all exams from October 18, 2020, to February 18, 2021, were included in the sampling process. They were distributed proportionally, with 755 used for training and validation, and 84 for testing, maintaining a balance between positive and negative cases The data sampling strategy considered the most restricted period for exam confidence.

As expected (see Table 11), adding data referring to tests with negative results performed outside the confidence interval (3 to 7 days from the onset of symptoms) does not improve the results. It highlights the high sensitivity and specificity for data within the range, reaching 76% and 82%, respectively, while data from negative tests outside this interval reached only 55% sensitivity and 70% specificity.

**Table 11.** Metrics data group 3 (RT-PCR 2*^nd^* wave) test set with and without the restriction of test reliability period RT-PCR - Random Forrest.

| Data group | Accuracy | Precision | Recall | f1 | AUC | Sensitivity | Specificity |
| --- | --- | --- | --- | --- | --- | --- | --- |
| 3 - with period restriction | 0.79 | 0.85 | 0.79 | 0.79 | 0.79 | 0.76 | 0.82 |
| 3 - without period restriction | 0.64 | 0.55 | 0.64 | 0.64 | 0.64 | 0.55 | 0.70 |

### Test with Data from a Different Wave

Usually, the training and validation of models are done with data before the test set so that it can be used to predict future results, but the exchange of test data between models trained with data from different times can help to identify the differences between the sets of data, aiming to evaluate the adherence of the behavior between the data. For this reason, data from the test set of the first symptomatic contagion wave of the virus SARS-CoV-2 were used in the model trained with data from the second wave, for example.

In order to change the test sets, it was necessary to fit the third wave test data set to the model trained with the first and second wave data. Thus, the attributes of back pain, ageusia, anosmia, and chest pain were added in the third wave test set (nasal congestion, sneezing, fatigue, joint pain, and nausea or vomiting were removed). On the other hand, when using the model trained with data from the third wave, it was necessary to adjust the test set of the first and second waves by removing the attributes low back pain, ageusia, anosmia, and chest pain and adding the attributes nausea or vomiting, joint pain, fatigue, sneezing and nasal congestion.

All these variables were available in the databases at the beginning of the attribute evaluation. Thus, seeking to increase the quality of the results obtained, some attributes were removed after the variable selection process. For this reason, the bases related to the 3*^rd^* wave had a distinct collection of characteristics from the 1*^st^* and 2*^nd^* waves.

Specifically, as the attributes of signs and symptoms between the first and second waves of contagion were the same, exchanging data from the two waves should produce close results. However, tests of models trained with data from a given wave and test sets from another were extended to all cases to evaluate all cases comprehensively.

Table 12 illustrates the results metrics, using the best model trained with data from the **first wave**, using different sets of tests. The decrease in sensitivity and recall when using a test set of other waves is noteworthy. In addition, in general, the results of the second wave test set outperform the results of the first wave tests, i.e., the test with data relative to the same set used for training, except for sensitivity and recall.

**Table 12.** Tests with RT-PCR 1*^st^* wave model (data group 2) - Random Forest.

| Data group | Accuracy | Precision | Recall | f1 | AUC | Sensitivity | Specificity |
| --- | --- | --- | --- | --- | --- | --- | --- |
| 2- RT-PCR 1<sup>st</sup> wave data | 0.65 | 0.66 | 0.65 | 0.65 | 0.65 | 0.62 | 0.69 |
| 3- RT-PCR 2<sup>nd</sup> wave data | 0.70 | 0.71 | 0.58 | 0.70 | 0.69 | 0.58 | 0.80 |
| 4- RT-antigens 3<sup>rd</sup> wave data | 0.61 | 0.61 | 0.40 | 0.61 | 0.59 | 0.40 | 0.78 |

As can be noticed in the table 13, the performance of the **second wave** model, using data from the test set of the wave itself, obtains excellent results. However, when tested with data from other waves, it starts to offer results below that what was obtained with the model trained with the first and third waves: the results of line 2 of Table 13 are below the results of line 1 of Table 12 and the results line 3 of Table 13 are also below the values obtained in line 1 of table 14, except in the latter case for the specificity metric. The decrease in sensitivity and recall to 38% with data from the 3*^rd^* wave is noteworthy.

**Table 13.** Tests with RT-PCR 2*^nd^* wave model (data group 3) - Random Forest.

| Data Group | Accuracy | Precision | Recall | f1 | AUC | Sensitivity | Specificity |
| --- | --- | --- | --- | --- | --- | --- | --- |
| 3 - RT-PCR 2<sup>nd</sup> wave data | 0.79 | 0.85 | 0.79 | 0.79 | 0.79 | 0.76 | 0.82 |
| 2 - RT-PCR 1<sup>st</sup> wave data | 0.62 | 0.61 | 0.60 | 0.62 | 0.62 | 0.60 | 0.65 |
| 4 - RT-antigens 3<sup>rd</sup> wave data | 0.62 | 0.64 | 0.38 | 0.62 | 0.60 | 0.38 | 0.81 |

**Table 14.** Tests with RT-antigens 3*^rd^* wave model (data group 4) - Random Forest.

| Data group | Acurácia | Precision | Recall | f1 | Área ROC | Sensibilidade | Especificidade |
| --- | --- | --- | --- | --- | --- | --- | --- |
| 4 - RT-antigens 3<sup>rd</sup> wave data | 0.72 | 0.72 | 0.70 | 0.72 | 0.71 | 0.70 | 0.73 |
| 2 - RT-PCR 1<sup>st</sup> wave data | 0.58 | 0.58 | 0.46 | 0.58 | 0.58 | 0.46 | 0.70 |
| 3 - RT-PCR 2<sup>nd</sup> wave data | 0.63 | 0.62 | 0.51 | 0.63 | 0.62 | 0.51 | 0.74 |

Likewise, Table 14 shows the tests of the **third wave** model with the test sets of the first and second wave separately. One can again observe the substantial worsening in almost all metrics, using data from different waves in the tests. Line 1 represents the tests using data from the third wave; it can be seen that all metrics were above 70%. On the other hand, in lines 2 and 3, which represent tests performed with data from the first and second wave, the metrics were much lower, except for specificity, which increased when data from the second wave was used in the model trained with data from the third. The sensitivity and recall of the model, which was 70%, with test data from the third wave, dropped to 46% using data from the first wave and 51% with data from the second wave. The models of the second wave and the third wave make it clear that training a model with data from one wave does not serve to predict test results of infection by SARS-CoV-2 on data from another wave; in other words, the models indicate that there are differences in signs and symptoms of SARS-CoV-2 infection between waves of symptomatic virus infection.

### Explainable

In order to add explanations for the results obtained,this subsection presents graphs of SHAP values of the SARS-CoV-2 infection predictors for the model RF for the RT-PCR bases 1*^st^*, 2*^nd^* and 3*^rd^* wave, using the test base of each wave. The choice of this subset of tests and databases can be justified because they were the subset with the best results.

Thus, the graphs in Figures 6,7 and 8 present the distribution SHAP of each record of their respective bases in relation to the performance of the models, where blue indicates low and red high values; positive red values indicate that the symptom’s presence helps the model identify it as positive for symptomatic infection by the SARS-CoV-2 virus.

**Fig 6.**
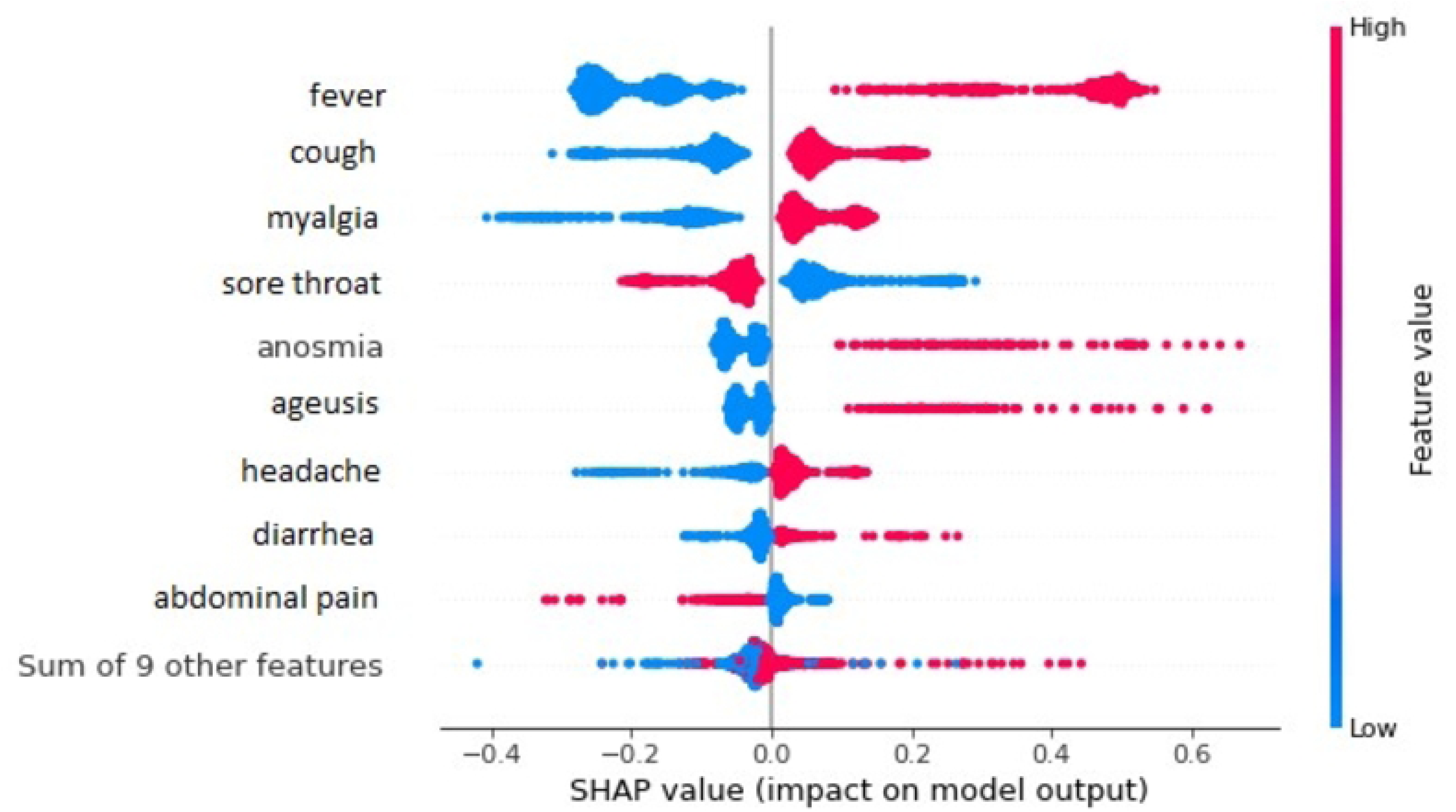

**Fig 7.**
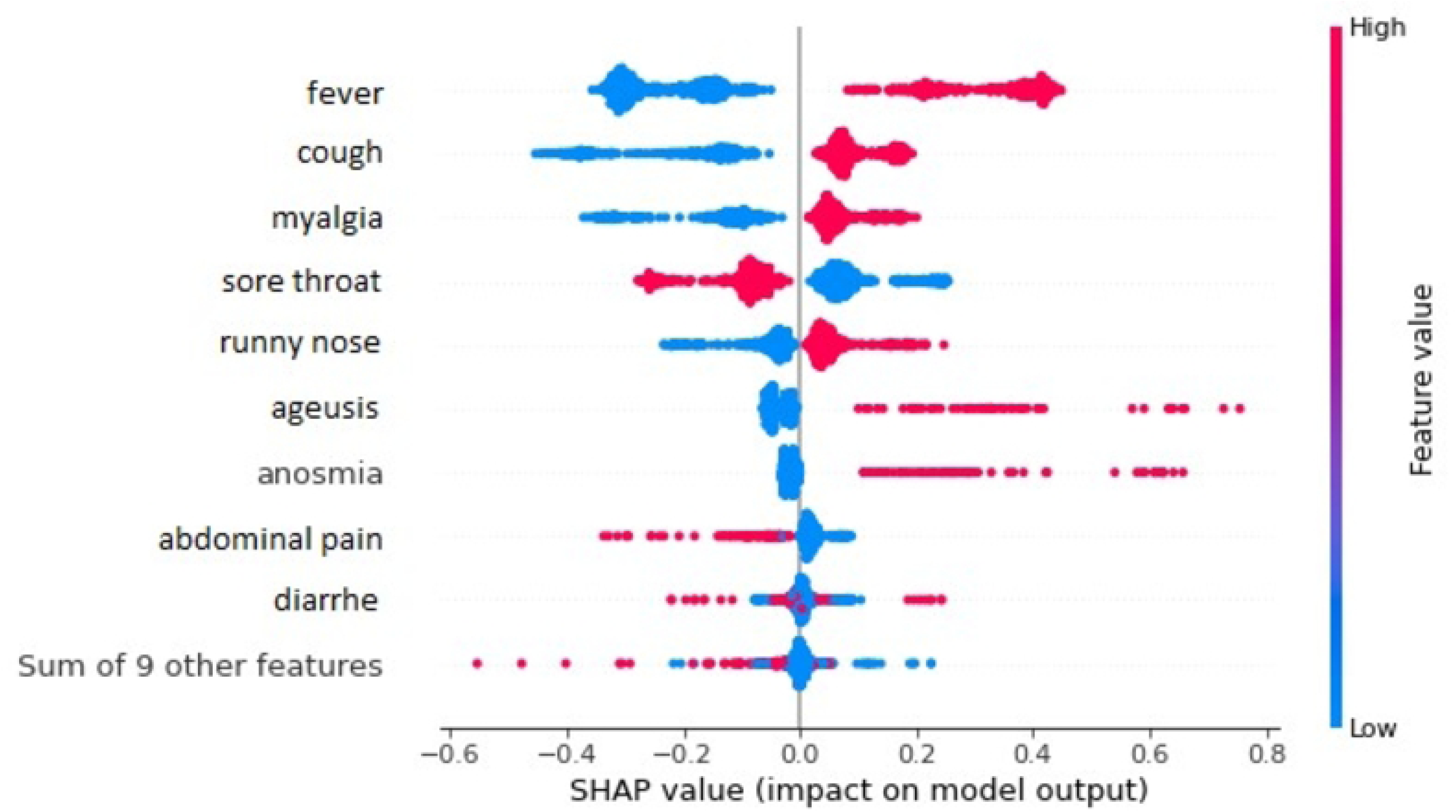

The graph illustrated in Figure 6 is formed by points that represent all the training data from the first wave of symptomatic infection by the virus SARS-CoV-2 for each attribute. As the base is binary, there can only be two colors in the graphs: blue must be 0 and red must be 1. The distribution of the points along the axis indicates greater or lesser impact for the positive (1) or negative (0), considering the attributes that are being classified. The occurrence of red and blue colors in opposite places indicates that these attributes are good predictors; after all, only by changing their value can the model verify its contribution to a class in a simpler way. These point clouds can also expand vertically, indicating that the density of values for that variable to SHAP values increases.

As can be seen, the attributes fever, cough and myalgia are the three most important signs and symptoms in explaining the prediction of infection in the 1*^st^* wave However, specifically having anosmia and ageusia indicated a high impact on classifying symptomatic infection by SARS-CoV-2, and not having one or the other did not contribute significantly to the classification quality. In addition, not having a sore throat or pain in the abdomen impacted the classification of SARS-CoV-2 infection.

Actually, not having a sore throat had a more significant impact on the indication of the absence of symptomatic infection by SARS-CoV-2.

Figure 7 presents the graph for SHAP values for the second wave of symptomatic infection by the virus SARS-CoV-2, where fever, cough, myalgia and sore throat had the most significant impact on the classification results, as did the first wave data. It should be noted that the coryza attribute becomes the most important 5*^th^* attribute for predicting infection by the virus, and the headache symptom no longer appears among the main ones, being together with eight other symptoms, represented by the last line from the chart.

In the graph indicated by Figure 8, it is observed that fever, the main symptom, remained in the graph, showing an even more significant impact. However, cough drops to the third position. Other attributes appear to be more important. Nasal congestion stands out, which was removed during the variable selection process for the second and first waves. Unlike previous waves, sneezing and asthenia are critical new attributes in the third wave.

**Fig 8.**
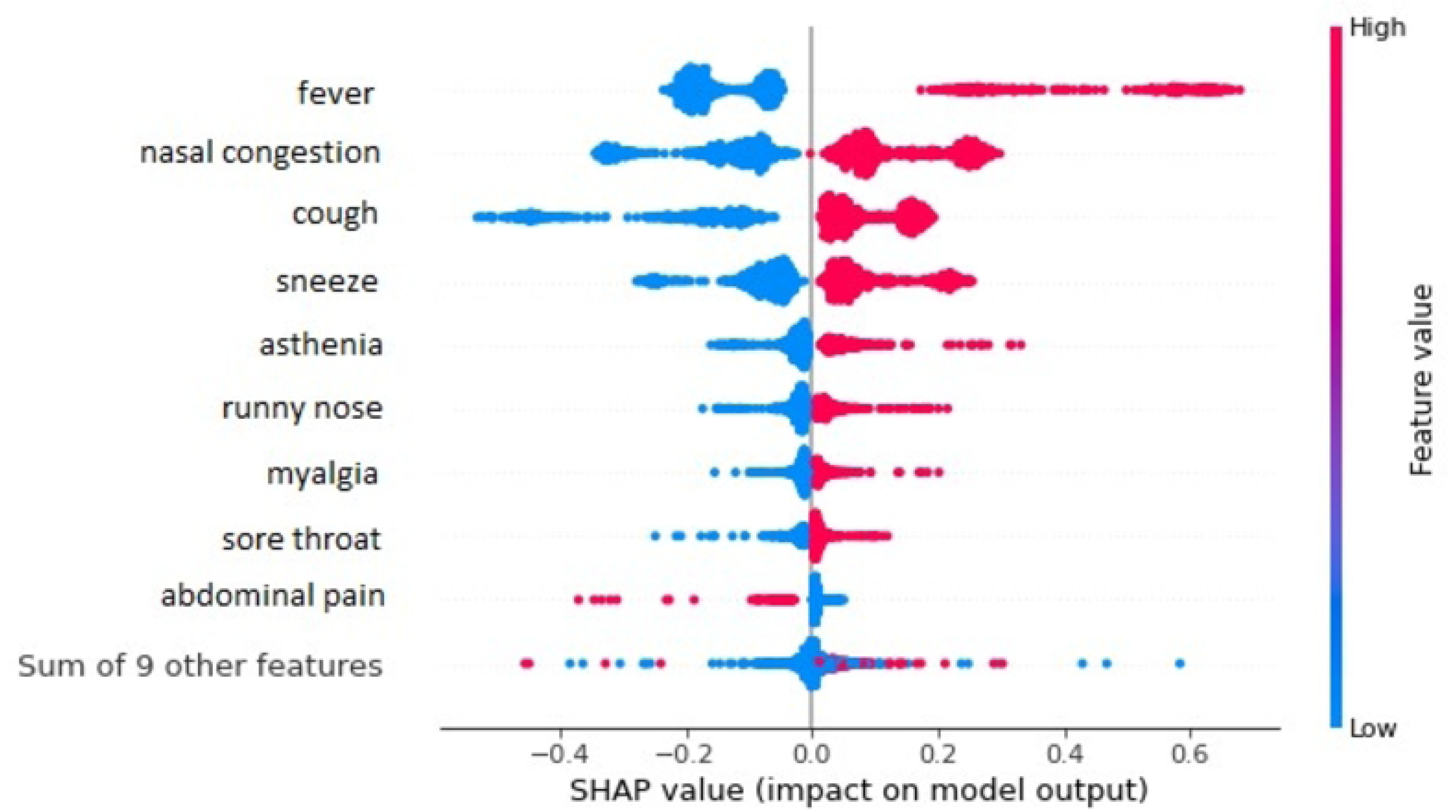

Figure 9 shows the explanation (generated by SHAP) that each attribute provides for predicting the diagnosis of a randomly chosen database record. This graph refers to a random value of the prediction of the RF model using the database RT-PCR first wave with the best validation result. The graph represents the impact of each attribute (sign or symptom) for predicting symptomatic SARS-CoV-2 virus infection for a specific input (a single patient); value closer to 1 is considered positive and closer to 0 is considered negative.

**Fig 9.**
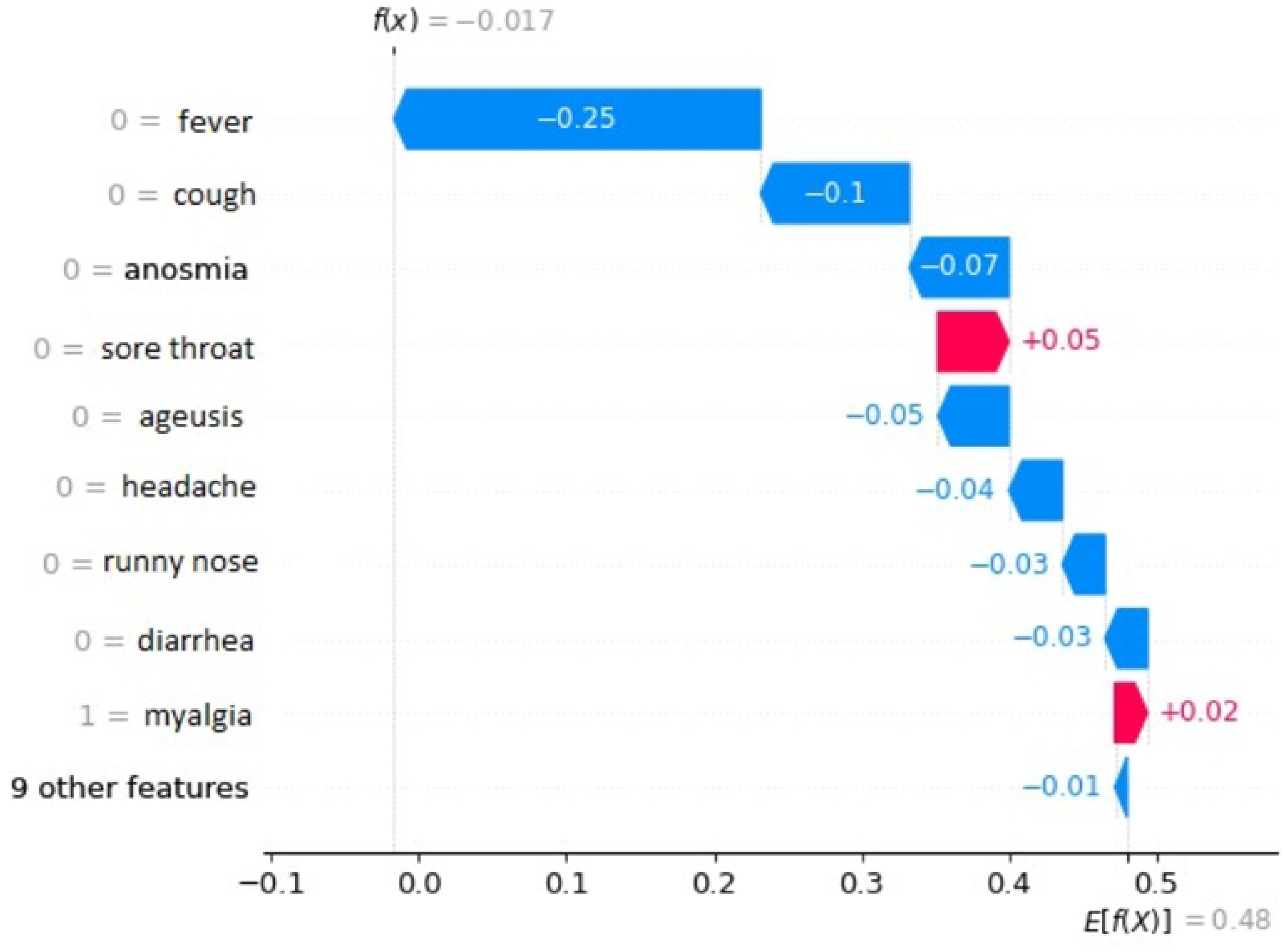

The visualization of the explanation produced by the Shapley method for this record which belongs to the group related to RT-PCR 2*^nd^* wave (group 3) with all the attributes described in Table 4, when evaluated by the RF model, can be seen in Figure 10. It can be seen that each attribute contributes positively (the model predicts a positive category) and negatively (the model predicts another class).

**Fig 10.**
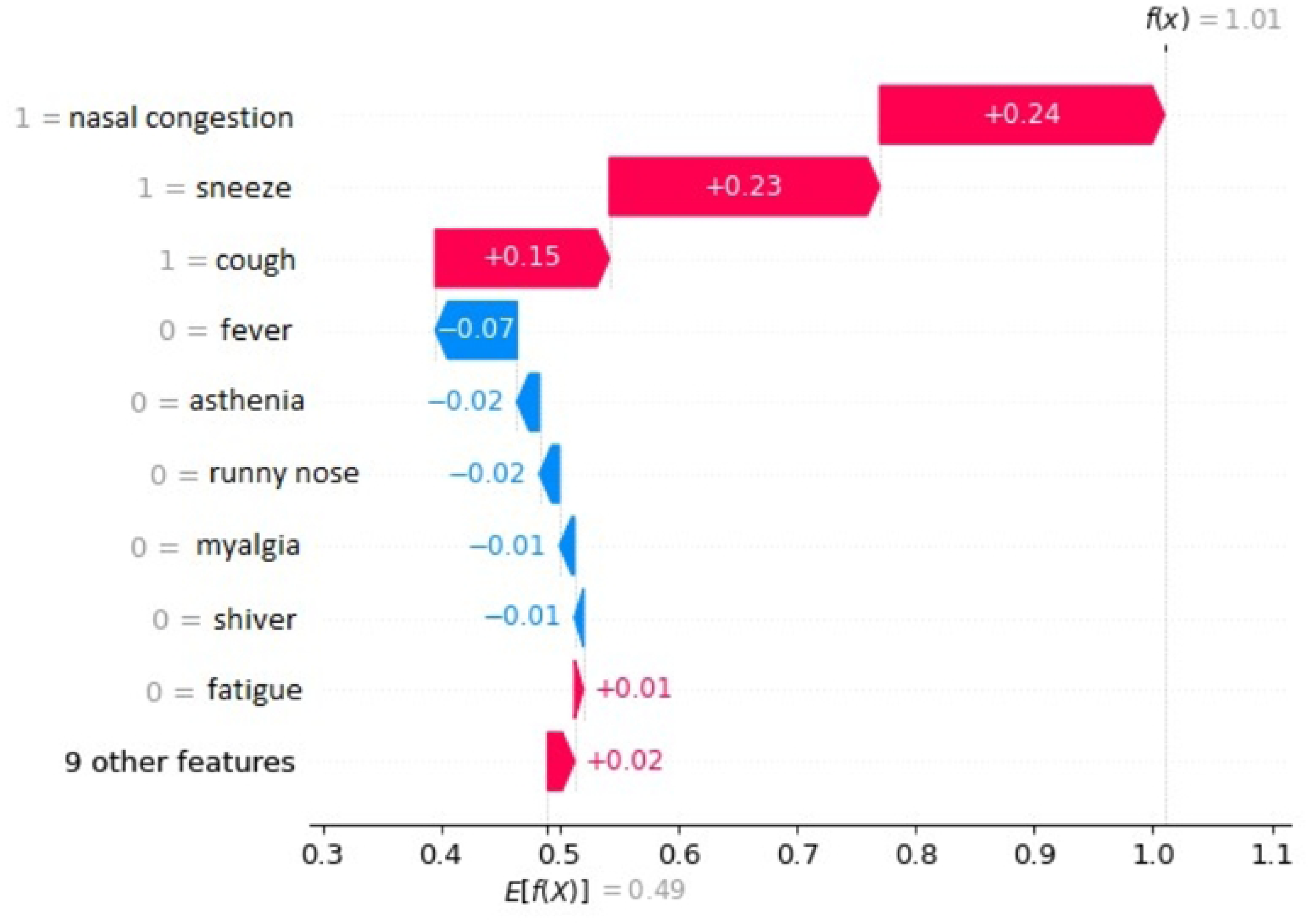

The X-axis represents the SHAP values and the arrow values indicate the contributions of these variables. Thus, fever and cough, on the graph, appear, indicating that the lack of these symptoms induces the model to classify as negative for symptomatic infection by SARS-CoV-2. On the other hand, the presence of a sore throat induces the model to classify as negative for symptomatic infection by SARS-CoV-2 in the 2*^nd^* wave.

Figure 11 displays the ratings for a specific patient. The graph indicates that the model predicted symptomatic infection by the virus SARS-CoV-2 from the reported signs and symptoms (attributes). The presence of nasal congestion, sneezing and coughing is a good indication for the model to be classified as positive for the virus. The lack of fever somewhat disturbed the model’s prediction.

**Fig 11.**
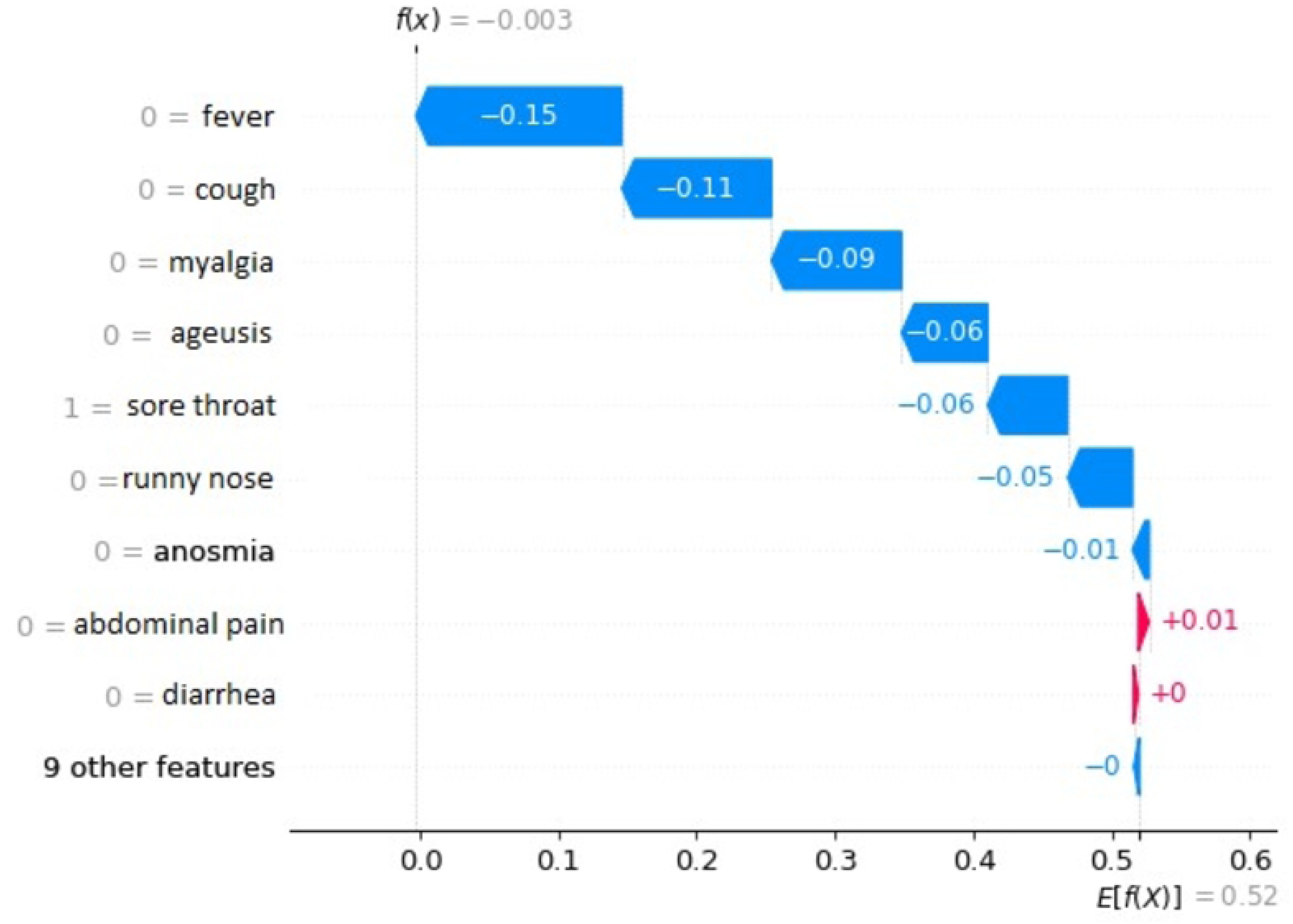

The scripts developed are registered under title “MultuModelosIA” and code BR512023000197-0, but are available for academic use.

## Discussion

Data were analyzed and divided into waves, through visual analysis of the moving average graph of confirmed cases of symptomatic infection by the SARS-CoV-2 virus.

In addition to signs and symptoms, other attributes were explored to improve the sensitivity and specificity of the models. The gender attribute was irrelevant for predicting the outcome of symptomatic infection with the SARS-CoV-2 virus. The information if the patient had contact with confirmed cases, markedly worsened the results. Analyzing the data from the third wave forms, it can be seen that many patients confirmed contact after a date after the onset of signs and symptoms, indicating that contact was not the cause of the infection. Data such as place of residence and age could not be used, as most patients did not fill in this information. In the database for the third wave, there was already information about the vaccine. However, as the vast majority of patients were vaccinated (97.5% vaccinated), it was not possible to use this information as an attribute to assess this base specifically.

Some Machine Learning models were used in exam databases that serve to detect the presence of symptomatic infection by the SARS-CoV-2 virus based on signs and symptoms only. The results of the tests used, which served as labels for the bases, were the RT-PCR test, the RT test of Antibodies and the RT test of Antigens, the latter only for the third wave. The first and second wave analyses used the RT Antibodies and RT-PCR tests.

It can be seen, through the results generated by the ML models, that the RT-PCR exam is much superior in detecting symptomatic infection by the SARS-CoV-2 virus; this is demonstrated by the metrics of the models when using the exams as labels separately. The best result of the model that considered the RT-PCR exam used data from the second wave, reaching 79% accuracy, 76% of sensitivity and 82% specificity, while the best model using the RT of Antibodies was 62% accuracy, with only 51% sensitivity and 67% specificity, considering the data from the first wave. All models that used data from the RT of Antibodies obtained lower results than the other models based on RT-PCR and RT of Antigens. The model’s best result using the RT of Antigens was 72% Accuracy with 70% sensitivity and 73% specificity, referring to the third wave.

The models of the first wave, in general, obtained worse results. The explanation for these results may lie in the fact that tests were scarce during the first wave, especially RT-PCR. At that time, the tests were reserved for people with comorbidities, the elderly and health professionals. In addition, in most cases, the tests were only performed on people with signs and symptoms; this fact may have led to false information on the part of patients.

Another aspect analyzed was the division into waves; the models were trained with data from the first and second waves and data outside a specific wave, that is, the period between waves. In this way, much more data is obtained; however, when mixing wave data, the models obtained inferior results; this fact motivated the testing of the models by training and validating them with data from a specific wave and testing with data from another wave. The cross-tests showed that the models trained in a particular wave do not obtain minimally acceptable results to predict the results of another wave We can highlight the model with data from the second wave that obtained excellent results, 79% accuracy, 76% sensitivity and 82% specificity; however, when tested with data from the third wave, it obtained only 62% accuracy with 38 % sensitivity and 81% specificity. These results show that models need to be trained with data from the wave itself. Another conclusion that can be drawn from this result is that the signs and symptoms that explain the prediction of symptomatic infection by the SARS-CoV-2 virus vary between waves.

Considering the previous tests and the signs and symptoms prevalence charts, an explainable technique was applied in order to assess the difference in these attributes between the different waves. It can be highlighted that between the first and second waves, there is a significant difference in the coryza attribute, which in the first wave did not appear among the nine main attributes; however, in the second wave, it appears as the 5*^th^* main symptom. Another attribute that changes from wave to wave in terms of importance for explaining infection prediction is headache, which does not appear as a top 9 in the second wave. The fever sign appears unchanged as being the most important for the explanation in the three waves; however, there are many changes, mainly in the third wave. It can be observed that ageusia and anosmia do not even appear among the top nine and nasal congestion, which disturbed the models in the previous waves, appears as the second most important attribute explaining the result. Notably, the sore throat attribute appeared as a symptom indicating the lack of infection in the first and second waves. Nevertheless, in the third wave, this attribute helps to classify as being positive for symptomatic infection by the SARS-CoV-2 virus.

## 5 Conclusion

This work aimed to develop a methodology for diagnosing symptomatic infection with the *SARS-CoV-2* virus to help health professionals triage patients and assist with social distancing. In addition, these models also allowed the study of the identification of the main signs and symptoms of each wave, the differences between the signs and symptoms observed in the waves of contagion of the *SARS-CoV-2* virus and the quality of the tests used in diagnosis. Considering the results obtained with the models, we believe that the objectives of this work have been achieved, with some models achieving values above 70% for the accuracy, sensitivity, and specificity metrics. From this model, it is understood that the methodology proved helpful in studying and evaluating the prevalence of signs and symptoms, considering the different waves of symptomatic infection by the *SARS-CoV-2* virus.

It can also be concluded that the models are viable for predicting symptomatic infection by the *SARS-CoV-2* virus, provided that data from the wave itself is used. This recommendation is based on the observation of the reduction in the values of the metrics when the test sets were exchanged between the waves. There is, therefore, evidence that the Omicron variant (3*^rd^* wave) is related to reports of different signs and symptoms. These models can help isolate patients more likely to be infected with the *SARS-CoV-2* virus and are quicker and cheaper than traditional tests. It was also concluded that *RT-PCR* and TR Antigen tests are considerably more reliable than TR antibody tests, as previously reported by Verotti [38].

As a future work, we intend to obtain new data on the variants that infected a given patient, including vaccines received, and thus identify which variant presents a given set of signs and symptoms. This same methodology could be evaluated for other diseases, such as *Dengue* or *Zika*. New attributes can also be included, containing other types of data, such as imaging or laboratory tests, using a multimodal approach or data fusion to investigate disease diagnoses and outcomes.

## Data Availability

All files are available from the OSF.io database. https://osf.io/d59g8/?view_only=6709b1471fe64cab8bc3208fec1e84b8

## Footnotes

1 Source: Municipal Health Secretariat of Rio de Janeiro - https://experience.arcgis.com/experience/38efc69787a346959c931568bd9e2cc4 - 11/17/2021

## Notes

### Competing Interest Statement

The authors have declared no competing interest.

